# Fetal Polygenic Growth Score and Risk for Large for Gestational Age Birth Weight in Nulliparas: Secondary Analysis of a Prospective Cohort Study

**DOI:** 10.1101/2025.10.28.25338986

**Authors:** Maha Aamir, Maisa Feghali, Lynn Yee, Robert M Silver, Celeste Durnwald, Jon G. Steller, David Haas, Rafael F Guerrero, Christina M Scifres

## Abstract

**Background:** Large for gestational age birth weight is associated with both short- and long-term health consequences for offspring, and fetal genetics may contribute to risk for large for gestational age birth weight.

**Objectives:** We evaluated the relationship between a polygenic growth score and the risk for large gestational age birth weight. We also delineated the between the polygenic growth score and risk for large for gestational age birth weight in relation to maternal glycemia and body mass index, both of which are established risk factors for large for gestational age birth weight.

**Study design:** This is a secondary analysis of a prospective multicenter cohort study in which nulliparous individuals were recruited from eight clinical sites in the United States. A subset of infants (n*=*3865) with DNA available were genotyped, and a previously developed polygenic growth score for large for gestational age birth weight was calculated. We evaluated the relationship between tertiles of the polygenic growth score, maternal body mass index, and glycemia assessed by the 50-gram glucose challenge test on the risk for large for gestational age birth weight using one-way ANOVA and Chi-squared tests as well as a regularized linear model.

**Results:** Of the 3,865 individuals with infant genotype available, 3,286 (84.9%) were included in this analysis. A polygenic growth score in the first tertile was associated with a lower risk for large for gestational age (OR 0.71, 95% CI 0.53-0.94), while a polygenic growth score in the third tertile was associated with a higher risk for large for gestational age birth weight (OR 1.29, 95% CI 1.02-1.63). Maternal body mass index was more strongly associated with the risk for large for gestational birth weight than maternal glycemia. The odds of large for gestational age birth weight were significantly higher with a maternal body mass index ≥35 kg/m^2^ and a PGS of either the second tertile (OR 3.54, 95% CI 1.96-6.38) or third tertile (OR 2.69, 95% CI 1.54-4.71).

**Conclusions:** The polygenic growth score has a modest ability to identify fetuses at higher or lower risk for Large for gestational age birth weight in a multiracial cohort from the United States. Polygenic growth score could assist with identification of those fetuses at increased risk for LGA birth weight among individuals with a BMI ≥35 kg/m^2^, which may allow for targeted interventions such as dietary and lifestyle modifications to optimize the in-utero environment.

## Introduction

Infants born large for gestational age (LGA), defined as a birth weight >90^th^ percentile, are at increased risk of adverse perinatal outcomes, increased mortality during the first year of life,^1,2^ and lifelong risk for obesity, dyslipidemia, hypertension, and type 2 diabetes.^2,3^ Both maternal diabetes and obesity are risk factors for LGA birth weight.^4,5^ Maternal glucose demonstrates a continuous association with offspring birth weight,^6^ with fetal hyperinsulinemia serving as a potent growth factor.^7^ However, maternal fasting glucose explains only 2-13% of the variance in birth weight,^8,9^ and the majority of infants born LGA are not born to mothers with diabetes.^10^ Rates of LGA are significantly increased among pregnant individuals with overweight or obesity both in the presence or absence of GDM.^5,11^

Genome-wide association study (GWAS) data have indicated an important role for maternal and fetal genetic variation in birth weight.^12-17^ Horikoshi, et al. identified robust associations of single nucleotide polymorphisms (SNPs) with birth weight at 60 individual loci in the fetal genome, some of which were known to be associated with glycemic traits in adults.^13^ Hughes, et al. used these data to create a weighted fetal genetic score for birth weight. This genetic score was associated with birth weight, independent of maternal fasting plasma glucose, in women of European ancestry and their offspring.^18^ Both fasting maternal plasma glucose and the fetal growth score were associated with increased fetal growth and had additive effects.^18^ However, they did not assess the impact of maternal body mass index (BMI) on fetal growth. We therefore aim to validate the relationship between a fetal genetic growth score, birth weight, and LGA birth weight in a large, multi-ancestry US cohort of nulliparous individuals. We also seek to further delineate the relationship between the fetal genetic growth score and the risk for fetal overgrowth in relation to maternal glycemia and BMI, both of which are established risk factors for LGA birth weight. We hypothesize that a combination of fetal genetic growth score and maternal factors will provide nuanced growth potential prediction and risk stratification during pregnancy.

## Materials and Methods

### Study population

This is a secondary analysis of the nuMoM2b (Nulliparous Pregnancy Outcomes Study: Monitoring Mothers-to-Be) Study. The protocol for the NuMoM2b study has been published previously.^19^ Briefly, the nuMoM2b study was a prospective multicenter cohort study in which nulliparous individuals were recruited from hospitals affiliated with 8 clinical sites in the United States (n=10,038). Each site’s local governing institutional review board approved the study, and all participants provided written informed consent before participation. Participants were enrolled from October 5, 2010, to December 3, 2013.

Among nuMoM2b with infant available biospecimens (n= 5,538), those who participated in the follow-up Heart Health Study were selected (n= 4,754). Participants were then filtered to retain the three largest genetic ancestry groups in the cohort (“EUR”, “AFR”, and “AMR”, assigned by genetic similarity to the European, African, and American superpopulations as defined in the 1K Genomes Project)^20^ to reduce the confounding effects of population stratification in our genetic analyses (n= 3994). We further excluded individuals with genetic data not available due to missing or insufficient blood specimens (n=86), those with low DNA yield so the PGS could not be calculated (n=40), those who failed mom-infant relatedness testing (n=7) and those with a genotyping rate <99% (n=5). In addition, we excluded those who did not have a glucose challenge testing during pregnancy (n=452), those with inferred and reported newborn sex mismatch (n=112) and those who were missing pregnancy outcome date or LGA status (n=6; Supplemental Figure 1). Our study followed the Strengthening the Reporting of Observational Studies in Epidemiology (STROBE) reporting guidelines.

### Covariates

Data were collected through interviews, self-administered questionnaires, clinical measurements, and medical records as part of the parent study. Pregnancy outcome information was collected from medical records, and maternal and cord blood samples were collected for DNA.^19^ Study visit 1 occurred between 6 0/7 and 13 6/7 weeks. At this time height and weight were measured, BMI (kg/m^2^) was calculated, and BMI was categorized into normal weight (BMI <25 kg/m2), overweight (BMI 25-29.9 kg/m^2^), and obese (BMI >=30 kg/m^2^). Gestational weight gain was calculated by first prenatal visit weight from delivery weight. Clinical information collected included maternal age, marital status, insurance type, household income, hypertension, pre-existing diabetes, and self-reported alcohol or tobacco use. Self-reported race and ethnicity were reported to describe the demographic characteristics of the cohort and assess generalizability. At least 30 days after delivery, trained and certified record abstractors reviewed the medical records of all participants and their neonates and recorded final birth outcomes.

Gestational diabetes (GDM) testing was performed per routine clinical care, with the majority of testing performed between 24-28 weeks using a 50 g one-hour glucose challenge test (GCT) followed by a 100g three-hour oral glucose tolerance test (OGTT) if the 50 g GCT was elevated. Gestational diabetes was diagnosed if 2 or more values on the 100g oral glucose tolerance test exceeded the following: 95 mg/dL fasting, 180 mg/dL at 1 hr., 155 mg/dL at 2 hrs., or 140 mg/dL at 3 hrs. The relationship between maternal glucose, the PGS, and LGA birth weight was assessed in two ways. First, we calculated tertiles of the 50g GCT. We next grouped individuals into 3 categories: normal glucose testing (either a 50 g GCT <130-140 mg/dL or a 50g GCT above this threshold with a 100g OGTT with no abnormal values), 1 value on the 100g OGTT that exceeded the Carpenter Coustan cut-offs, or GDM diagnosed with 2 or more abnormal values using the Carpenter Coustan Criteria.

### Genetic data processing

Infants were genotyped infants (n*=*3865) using the Infinium Multi-Ethnic Global D2 BeadChip (1,748,280 markers; Illumina, Miami, USA), and maternal genotype was collected previously (n= 9757).^21^ Raw intensity data (.idat files) were inspected and called for the loci that passed initial quality control (97% of all markers in the array) using Beeline autoconvert (Ilumina).

These files were then converted to Variant Call Format (VCF) using the gtc2vcf plugin from bcftools.^22^ Once obtained, the vcf files were processed to only retain single nucleotide variants with genotyping rate > 99%, minor allele frequency > 0.01, and that were under Hardy-Weinberg equilibrium (i.e. HWE P > 5×10^−2^). KING 2.2.7^23^ was used to confirm genetic relatedness between mothers and infants, filtering out pairs that showed very high relatedness suggesting either contamination or duplication of the samples (kinship coefficient > 0.354) (n= 14). The remaining samples (n= 3853) were phased with EAGLE and imputed via the TOPMED Imputation Server (R2).^24^ The genetic sex of infants was inferred and samples where the reported phenotypic sex differed from inferred genetic sex were removed (112 cases; Supplemental Figure 1).

### Study Variables

Our primary outcome was birth weight, which was evaluated as a continuous variable and a categorical variable. Birth weight was adjusted for infant sex and gestational age. Small for gestational age (<10^th^ percentile) and large for gestational age (>10^th^ percentile) were defined using a United States birth weight reference corrected for implausible gestational age estimates and assigned based on newborn sex.^25^

Maternal outcomes that may be related to birth weight were assessed including cesarean delivery, gestational diabetes, and hypertensive disorders of pregnancy (HDP). HDP were defined as preeclampsia/eclampsia (including preeclampsia superimposed on chronic hypertension) or gestational hypertension. HDP diagnoses were ascertained through 14 days postpartum by manual chart abstraction.^19^

A polygenic growth score (PGS) for birth weight that was validated in previous research was calculated for the infant genotype via PLINKv1.9.^18,26^ This PGS, which consisted of 60 variants, was developed from a cohort of European genetic ancestry. To allow for the application of this EUR PGS on our admixed cohort (i.e., to control for population stratification), this raw score was then further adjusted via the first ten principal components (linear model PRSraw ∼ PC1+PC2+PC3 …+PC10) in R (v.4.0.3).^27^ We used tertiles of the distribution of this adjusted score as a categorical variable in downstream analyses. The final cohort that was used for our analysis contained mother-infant dyads (n= 3286). As an exploratory analysis, we also assessed the impact of applying the PGS to the maternal genotype to assess the relationship between the PGS and birth weight.

### Statistical modelling for maternal and fetal polygenic growth scores

First, we explored correlates of LGA by one-way ANOVA and Chi-squared test, for continuous and categorical variables, respectively. Then, we developed a regularized linear model (Elastic Net^28^) of LGA from genetic and phenotypic data, including PGS alongside BMI and the 50 g GCT results, as well as all pairwise interaction effects among these three main factors.

## Results

LGA birth weight occurred in 257 (7.8%) of the 3,286 individuals included in this analysis, as shown in Table 1, maternal demographic characteristics including age, race, insurance status, education, BMI (both as a continuous and categorical variable), and chronic hypertension were similar among groups. There were differences in the number of individuals who reported any tobacco use during pregnancy across PGS tertiles (14.14 vs 19.00 vs 17.44%, p<0.001; Table 1). In addition, there was a trend towards slightly higher weight gain with increasing PGS tertile (Table 1). There were similar results on the GCT among groups, and no differences in the prevalence of mild glucose intolerance or GDM among groups.

**Table 1.** Maternal demographics and glycemia by polygenic growth score tertile using offspring genome.

| Characteristic | Growth Score Tertile 1 (n=1096) | Growth Score Tertile 2 (n=1095) | Growth Score Tertile 3 (n=1095) | P |
| --- | --- | --- | --- | --- |
| Maternal age (years) | 26.83 ± 5.4 | 26.33 ± 5.3 | 26.83 ± 5.4 | 0.977 |
| Race |  |  |  | 0.402 |
| White | 731 (66.7) | 712 (65.0) | 753 (68.8) |  |
| Black | 144 (13.1) | 145 (13.2) | 124 (11.3) |  |
| Hispanic | 166 (15.2) | 183 (16.7) | 171 (15.6) |  |
| Education |  |  |  | 0.053 |
| High school or less | 184 (16.8) | 226 (20.6) | 218 (19.9) |  |
| At least some college | 910 (83.0) | 868 (79.3) | 876 (80.0) |  |
| Insurance |  |  |  | 0.051 |
| Private | 804 (73.4) | 753 (68.8) | 810 (74.0) |  |
| Public | 271 (24.7) | 323 (29.5) | 269 (24.6) |  |
| Other | 13 (1.2) | 11 (1.0) | 10 (0.9) |  |
| Mean pre-pregnancy BMI (kg/m<sup>2</sup>) | 26.64 ± 6.3 | 26.53 ± 6.3 | 26.74 ± 6.6 | 0.733 |
| Pre-pregnancy BMI category |  |  |  | 0.454 |
| Underweight | 20 (1.8) | 17 (1.6) | 26 (2.34) |  |
| Normal weight | 514 (46.9) | 541 (49.4) | 540 (49.3) |  |
| Overweight | 286 (26.1) | 248 (22.7) | 260 (23.7) |  |
| Obese | 254 (23.2) | 262 (23.9) | 259 (23.7) |  |
| Weight gain (lbs) | 33.77 ± 16.7 | 34.66 ± 15.9 | 35.18 ± 17.2 | 0.05 |
| Chronic hypertension | 19 (1.7) | 27 (2.5) | 31 (2.8) | 0.224 |
| Tobacco use | 155 (14.1) | 208 (19.0) | 191 (17.4) | 8.2 x 10<sup>-3</sup> |
| GDM testing (weeks) | 26.57 ± 7.0 | 26.49 ± 7.0 | 26.84 ± 5.6 | 0.343 |
| Mean 50 g GCT result (mg/dL) | 110.19 ± 27.7 | 110.86 ± 31.0 | 110.57 ± 27.7 | 0.754 |
| Mild glucose intolerance* | 30 (2.7) | 27 (2.5) | 26 (2.4) | 0.853 |
| GDM | 45 (4.1) | 54 (4.9) | 48 (4.4) | 0.636 |

Legend: Data shown as n (%) or mean ±SD. BMI (body mass index); GCT (Glucose challenge test); GDM (gestational diabetes). \*Mild glucose intolerance was defined as one abnormal value on the 100g OGTT.

The fetal PGS was associated with birth weight, with a mean difference of 68 g between PGS tertiles 1 and 2 and a mean difference of 65 g between GS tertiles 2 and 3 (ANOVA *P*=3.32 × 10^−14^; Table 2). The prevalence of LGA birth weight increased and small for gestational age (SGA) birth weight decreased as the PGS tertile increased (Table 2). There were no differences in cesarean delivery or HDP across PGS tertiles. We next conducted stratified analyses evaluating mean birth weight and LGA birth weight across 50g GCT tertiles, GDM diagnosis categories, and BMI categories (Table 3). There was a statistically significant increase in adjusted mean birth weight by PGS categories across 50g GCT categories and BMI categories. There was also an increase in mean birth weight by PGS tertile among those with normal GDM testing and with GDM (Table 3). There was a statistically significant difference in rates of LGA birth weight among those with a GCT value <97 mg/dL (4.4 vs 4.5 vs 8%, p=5.98x 10^−2^) and GCT between 97-118 mg/dL (6.0 vs 9.9 vs 10.8%, p= 5.82x 10^−2^). In addition, the prevalence of LGA birth weight differed significantly among those with a BMI <25 kg/m^2^ (4.1 vs 5.9 vs 8.0%, p=0.028).

**Table 2.** Pregnancy outcomes by polygenic growth score tertile using offspring genome.

| **Outcome** | **Growth Score**<br>**Tertile 1**<br>**(n=1096)** | **Growth Score**<br>**Tertile 2**<br>**(n=1095)** | **Growth Score**<br>**Tertile 3**<br>**(n=1095)** | **P** |
| --- | --- | --- | --- | --- |
| Gestational age at delivery (weeks) | 38.9 ± 1.7 | 38.8 ± 1.8 | 39.0 ± 1.6 | 0.766 |
| Preterm birth | 84 (7.7) | 87 (8.0) | 81 (7.4) | 0.89 |
| Birth weight (g) | 3231 ± 400 | 3299 ± 406 | 3364 ± 423 | 3.32 × 10<sup>-14</sup> |
| Birth weight category |  |  |  |  |
| AGA | 973 (88.8) | 960 (87.7) | 954 (87.1) | 3.36 × 10<sup>-4</sup> |
| SGA | 61 (5.6) | 48 (4.4) | 33 (3.0) |  |
| LGA | 62 (5.7) | 87 (8.0) | 108 (10.0) |  |
| Neonatal sex |  |  |  |  |
| Female | 545 (49.7) | 537 (49.0) | 561 (51.2) | 0.577 |
| Male | 551 (50.3) | 558 (51.0) | 534 (48.8) |  |
| Mode of delivery |  |  |  |  |
| Vaginal | 835 (76.2) | 821 (75.0) | 792 (72.3) | 0.104 |
| Cesarean | 261 (23.8) | 273 (24.9) | 303 (27.7) |  |
| Hypertensive disorders of pregnancy | 295 (26.9) | 311 (28.4) | 301 (27.5) | 0.727 |

Legend: Data shown as n (%) or mean ±SD. AGA (Appropriate for gestational age); SGA (small for gestational age); LGA (large for gestational age)

**Table 3.**
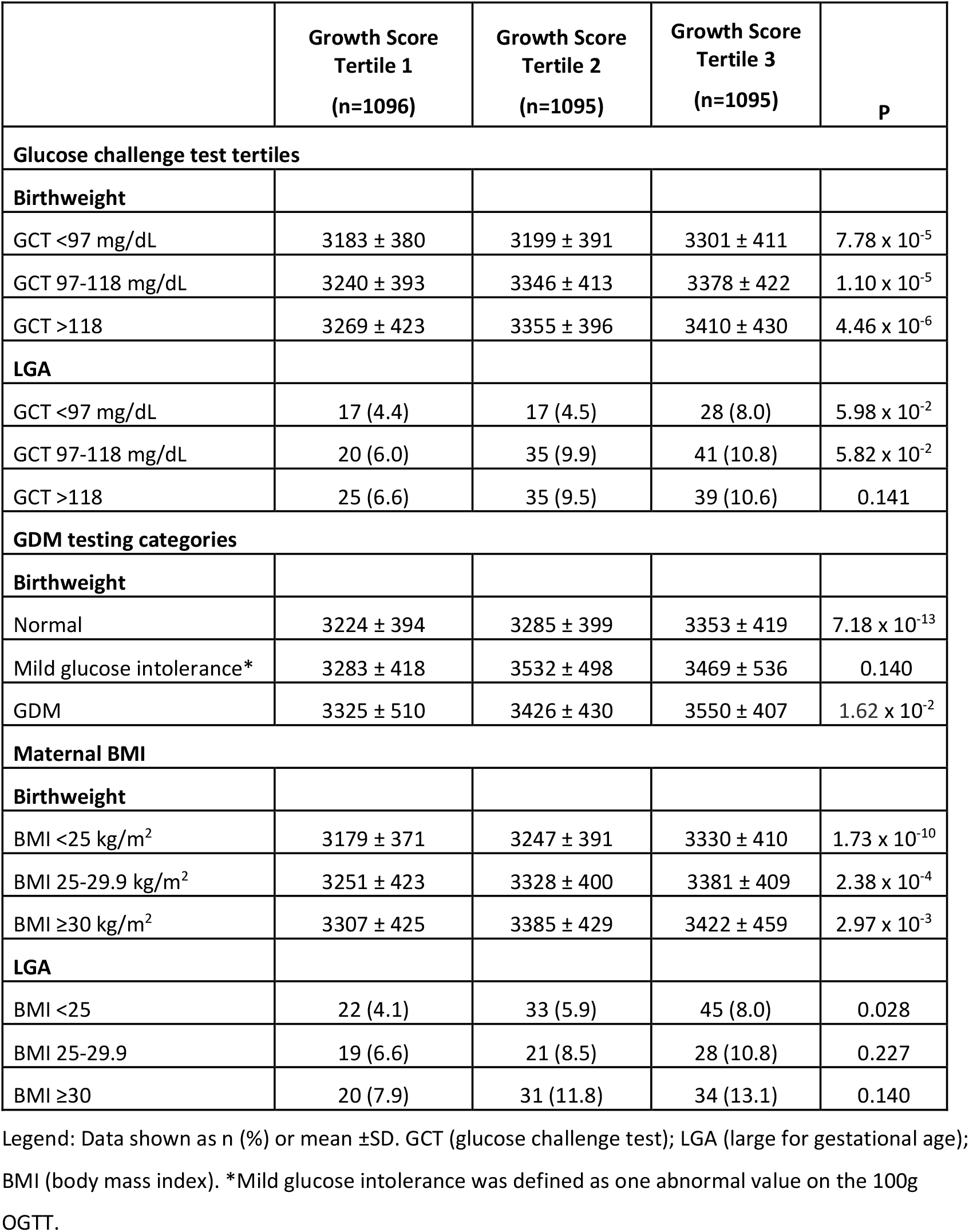
Relationship between the polygenic growth score tertile using the offspring genome, maternal glycemia, and maternal BMI.

|  | Growth Score<br>Tertile 1<br>(n=1096) | Growth Score<br>Tertile 2<br>(n=1095) | Growth Score<br>Tertile 3<br>(n=1095) | P |
| --- | --- | --- | --- | --- |
| **Glucose challenge test tertiles** |  |  |  |  |
| **Birthweight** |  |  |  |  |
| GCT <97 mg/dL | 3183 ± 380 | 3199 ± 391 | 3301 ± 411 | 7.78 × 10<sup>-5</sup> |
| GCT 97-118 mg/dL | 3240 ± 393 | 3346 ± 413 | 3378 ± 422 | 1.10 × 10<sup>-5</sup> |
| GCT >118 | 3269 ± 423 | 3355 ± 396 | 3410 ± 430 | 4.46 × 10<sup>-6</sup> |
| **LGA** |  |  |  |  |
| GCT <97 mg/dL | 17 (4.4) | 17 (4.5) | 28 (8.0) | 5.98 × 10<sup>-2</sup> |
| GCT 97-118 mg/dL | 20 (6.0) | 35 (9.9) | 41 (10.8) | 5.82 × 10<sup>-2</sup> |
| GCT >118 | 25 (6.6) | 35 (9.5) | 39 (10.6) | 0.141 |
| **GDM testing categories** |  |  |  |  |
| **Birthweight** |  |  |  |  |
| Normal | 3224 ± 394 | 3285 ± 399 | 3353 ± 419 | 7.18 × 10<sup>-13</sup> |
| Mild glucose intolerance* | 3283 ± 418 | 3532 ± 498 | 3469 ± 536 | 0.140 |
| GDM | 3325 ± 510 | 3426 ± 430 | 3550 ± 407 | 1.62 × 10<sup>-2</sup> |
| **Maternal BMI** |  |  |  |  |
| **Birthweight** |  |  |  |  |
| BMI <25 kg/m<sup>2</sup> | 3179 ± 371 | 3247 ± 391 | 3330 ± 410 | 1.73 × 10<sup>-10</sup> |
| BMI 25-29.9 kg/m<sup>2</sup> | 3251 ± 423 | 3328 ± 400 | 3381 ± 409 | 2.38 × 10<sup>-4</sup> |
| BMI ≥30 kg/m<sup>2</sup> | 3307 ± 425 | 3385 ± 429 | 3422 ± 459 | 2.97 × 10<sup>-3</sup> |
| **LGA** |  |  |  |  |
| BMI <25 | 22 (4.1) | 33 (5.9) | 45 (8.0) | 0.028 |
| BMI 25-29.9 | 19 (6.6) | 21 (8.5) | 28 (10.8) | 0.227 |
| BMI ≥30 | 20 (7.9) | 31 (11.8) | 34 (13.1) | 0.140 |

Legend: Data shown as n (%) or mean ±SD. GCT (glucose challenge test); LGA (large for gestational age); BMI (body mass index). \*Mild glucose intolerance was defined as one abnormal value on the 100g OGTT.

Modeling was conducted to assess the independent effects of the PGS on LGA while accounting for maternal glycemia and BMI. A modest and synergistic interaction between BMI and the fetal genetic score was identified. Figure 1 demonstrates that a PGS in the 1^st^ tertile was associated with a lower risk for LGA (OR 0.71, 95% CI 0.53-0.94), while a PGS in the 3^rd^ tertile was associated with higher risk for LGA (OR 1.29, 95% CI 1.02-1.63) when compared to the population-based LGA prevalence. Maternal BMI ≥35 kg/m^2^ was associated with a higher risk for LGA (OR 2.16, 95% CI 1.58-2.95), while a BMI <25 kg/m^2^ was associated with a reduced risk for LGA birth weight (OR 0.76, 95% CI 0.60-0.96). Higher values on maternal GCT were not associated with an increased risk for LGA birth weight, but a GCT <97 mg/dL was associated with a lower risk. Ris for LGA birth weight were significantly higher with a maternal BMI ≥35 kg/m^2^ and a PGS of either the 2^nd^ tertile (OR 3.54, 95% CI 1.96-6.38) or 3^rd^ tertile (OR 2.69, 95% CI 1.54-4.71), while the risk for LGA in the 1^st^ tertile was similar to the population-based prevalence of LGA.

**Figure 1.**
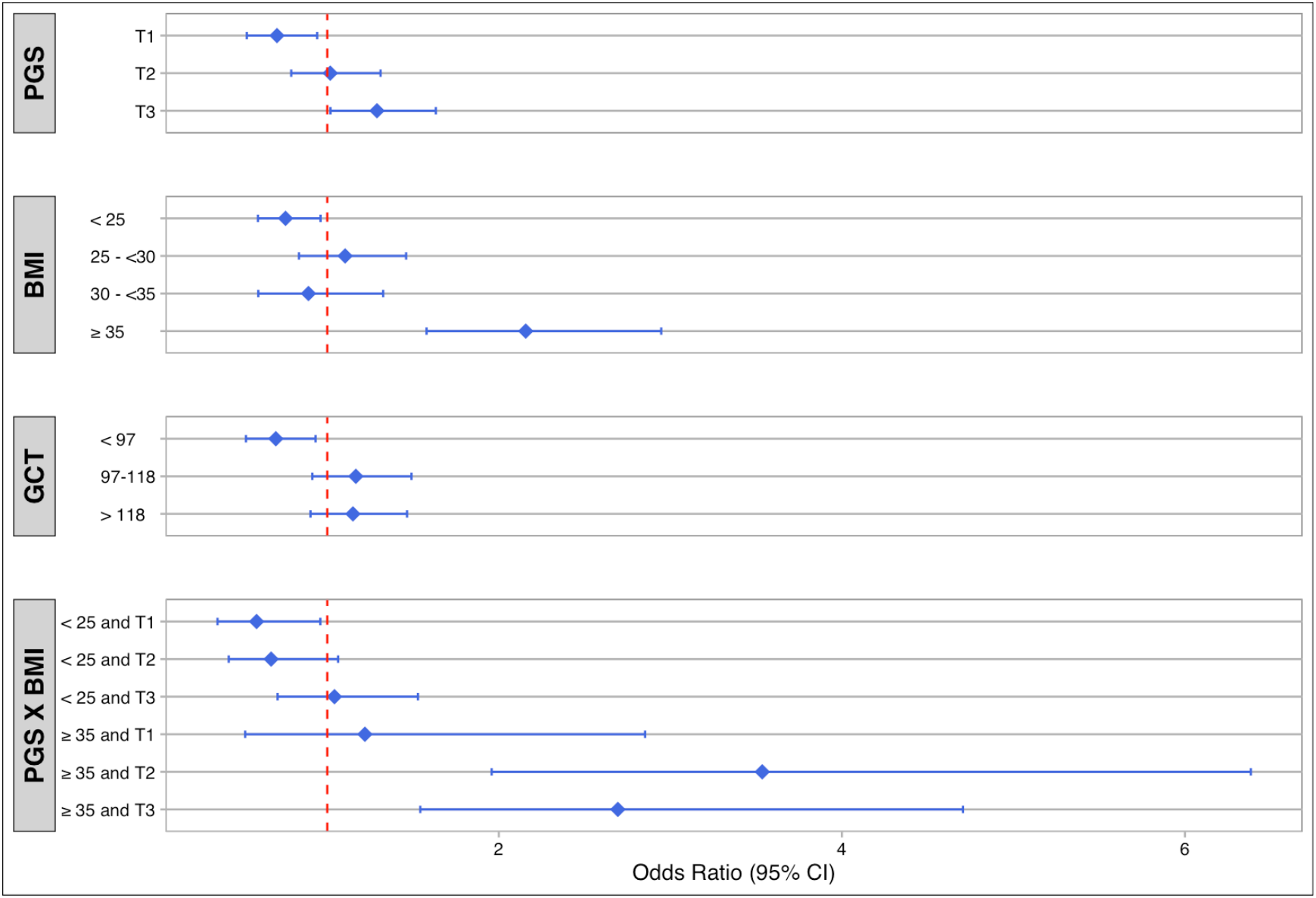
Odds ratios for LGA risk of predictors identified by the model. The odds ratios are defined with respect to the odds of LGA in the general nuMom2b population. Polygenic growth score (PGS), body Mass Index (BMI), glucose challenge test (GCT), results of interaction testing between the PGS and BMI (PGS x BMI)

When considered alone, the PGS had limited ability to identify LGA risk (AUC 0.58; 95% CI: 0.54-0.62 (*DeLong*; Supplemental Figure 3A). The AUC values improved somewhat with the addition of maternal BMI and 50g GCT values (AUC 0.64; Supplemental Figure 3B), and the fully adjusted model which included maternal BMI, 50 g GCT values, chronic hypertension, smoking, and maternal weight gain (AUC 0.69, Supplemental Figure 3C). In this model, the strongest predictors of LGA were maternal early pregnancy BMI (Beta=0.31) and fetal growth score (Beta=0.19), while the 50 g GCT was not significant.

As an exploratory analysis, we next utilized the PGS against the maternal genome to assess PGS performance (n=7843) given the genetic similarities between mother and child and the increased accessibility of the maternal genome. Supplementary tables 1 and 2, indicated that the results for the PGS used on the maternal genome were overall similar to those from the newborn genome. Both the mean birth weight and the likelihood of LGA birth weight increased across PGS tertiles. When stratified by maternal 50 g GCT results and BMI, the results were similar to results using the infant genotype. The AUC for the models adjusted for BMI and 50 g GCT had similar values for the newborn and maternal genetic scores (0.64 vs 0.61, p=0.30; Supplementary Figure 3B), as did the fully adjusted models (0.69 vs 0.66; p=0.06).

## Comment

### Principal Findings

A PGS has a modest ability to identify fetuses at higher or lower risk for LGA in a multiracial cohort from the United States. When compared to the overall population, a PGS in the highest tertile was associated with an increased risk for LGA birth weight while a PGS score in the lowest tertile was associated with a reduced risk for LGA birth weight. There also was evidence of an interaction between PGS and maternal BMI. Specifically, among individuals with a BMI ≥35 kg/m^2^, a PGS in the 1^st^ tertile was associated with lower risk of LGA birth weight, while a PGS score in the 2^nd^ and 3^rd^ tertiles identified a subset of fetuses at elevated risk for LGA birth weight. Although the AUC for the PGS alone indicates a limited ability to predict LGA birth weight, our results suggest that the PGS could assist with identification of those fetuses at increased risk for LGA birth weight among individuals with a BMI ≥35 kg/m^2^, which may allow for targeted interventions such as dietary and lifestyle modifications to optimize the in-utero environment.

### Results in the Context of What is Known

Our results highlight the complex relationship between maternal obesity, glucose, and LGA birth weight. Catalano, et al. found that both obesity and GDM are related to risk for LGA birth weight, and together have an additive effect on this risk.^29^ We found that maternal BMI was more strongly associated with the risk for LGA birth weight than the 50 g GCT results, and we identified a significant interaction only between maternal BMI and the PGS. This finding is in contrast to the finding of Hughes, et al., who assessed the relationship between fetal genetic growth score tertiles and fasting glucose and found that the risk for LGA birth weight ranged from 13.3% to 31.1% across growth score tertiles in the highest fasting glucose category.^18,30^ However, they did not consider maternal obesity in their analyses and maternal hyperglycemia was untreated in their cohort.

The mechanisms by which genetic risk may contribute to birth weight are also important. Hughes, et al. previously found that neither cord insulin nor cord C-peptide were associated with the fetal genetic score, indicating that the collective mechanisms of action of the SNPs in the fetal genetic score are largely independent of fetal insulin secretion.^18^ Work by Juliusdottir, et al. evaluated parental and fetal contributions to birth weight in an Icelandic cohort. They found that there is a complex pattern of inheritance affecting fetal growth. Fetal genetics appeared to have the strongest association with birth weight, while the maternal genome contributed to birth weight through variants associated with glycemic traits.^15^

### Clinical Implications

Prediction of LGA birth is difficult despite known risk factors for LGA birth weight including height, parity, ethnicity, age, prior delivery of an LGA infant, and fetal sex ^31,32^ and modifiable risk factors including maternal BMI and gestational weight gain.^6,31,33^ A genetic score that could further refine these risks would be welcome. Chawla, et al. explored the relationship between SNPs in 40 regions associated with adult obesity-related traits and tested for their association with newborn size. They identified 25 and 23 SNPs that were associated with birth weight and newborn adiposity. Addition of this genetic risk score to a model including known risk factors improved prediction of birth weight >90^th^ percentile and sum of skin folds >90^th^ percentile in four ancestry groups.^34^ While ultrasound can help identify higher birth weight, it provides only modest predictive ability for newborn adiposity.^35^

### Research Implications

Progress is being made in whole-genome sequencing of cell-free fetal DNA,^37-40^ and techniques to detect multiple SNPs associated with a complex trait such as birth weight may be available in the future. This may add even more nuance to our ability to predict LGA birth weight and guide timely interventions aimed at optimizing the in-utero environment for fetal growth and development. In addition, our finding that the fetal growth score performed similarly when using the maternal genome is another avenue in which genetic evaluation using maternal samples may be able to help shed light on risks for the developing fetus.

### Strengths and Limitations

Strengths of our study include the large sample size and robust characterization of the cohort including precision in gestational dating, which is critical for accurate determination of fetal growth. Only nulliparous individuals were included, which limits possible confounding related to parity. One limitation is that we did not have information on newborn body composition. The adverse metabolic consequences associated with LGA are likely secondary to excess fat as opposed to lean body mass;^36^ therefore identification of risk factors for newborn adiposity is a key challenge. We also did not include information on maternal nutrition, which may have an impact on fetal growth and development.

## Conclusions

A polygenic growth score could assist with identification of those fetuses at increased risk for LGA birth weight among individuals with a BMI ≥35 kg/m^2^, which may allow for targeted interventions such as dietary and lifestyle modifications to optimize the in-utero environment.

## Data Availability

All data produced in the present study are available upon reasonable request to the authors. Data will be uploaded to dBGap upon publication

